# INTRAREGIONAL PROPAGATION OF COVID-19 CASES IN PARÁ, BRAZIL. ASSESSMENT OF ISOLATION REGIME TO LOCKDOWN

**DOI:** 10.1101/2020.06.10.20127886

**Authors:** Félix Lélis da Silva, Javier Dias Pita, Maryjane Diniz A. Gomes, Andréa P. Lélis da Silva, Gabriel Lélis P. da Silva

## Abstract

Due to the high incidence of COVID-19 case numbers internationally, the World Health Organization (WHO) declared a Public Health Emergency of global relevance, advising countries to follow protocols to combat pandemic advance through actions that can reduce spread and consequently avoid a collapse in the local health system. On March 18, 2020, Pará notified the first case of COVID-19. After seven weeks, the number of confirmed cases reached 4,756 with 375 deaths. Knowing that infected people may be asymptomatic, the disease symptomatology absence and the population’s neglect of isolation influence the spread, and factors such as chronic pneumonia, high age, obesity, chronic kidney diseases and other comorbidities favor the mortality rate. On the other hand, social isolation, quarantine and lockdown seek to contain the intraregional contagion advance. This study analyzes the dynamics of COVID-19 new cases advance among municipalities in the state of Pará, Brazil. The results show it took 49 days for 81% of the state’s municipalities to register COVID-19 cases. The association between social isolation, quarantine and lockdown as an action to contain the infection was effective in reducing the region’s new cases registration of COVID-19 in the short-term.

## 1. Introduction

The COVID-19 pandemic stands out as the main global health crisis (Wu et al., 2020). It started in Wuhan, China in December 2019 (Şahin, 2020; Ahmadi et al., 2020; See et al., 2020). It is a respiratory infection caused by the coronaviruses family (2019-nCoV) (Prata et al., 2020). Its etiological agent is Sars-CoV-2 (Saez et al., 2020; Yang et al., 2020), which causes severe acute respiratory syndrome (SARS) (Solé at al., 2020). The viral infection presents severe clinical symptoms such as fever, dry cough, dyspnea, and pneumonia (Coccia, 2020; Wu et al., 2020) and can cause the death of the infected (Waldecy; David; Wainesten; 2020). Flu is one of the main causes of illnesses and death in the world (Mertz et al., 2013).

Epidemiological data associated with COVID-19 infection has shown different dynamics in several countries (Marson; Ortega, 2020). Several studies have shown the reverse effect of rising temperatures and the confirmed number of new cases (Zhu and Xie, 2020; Wu et al. 2020). Brazil, a country with a tropical climate, with an average annual temperature ranging from 16 to 27.4 °C, modeled results show a negative effect on the linear relationship of temperature with the new confirmed cases number (Prata et al., 2020).

Several factors have contributed to pandemic advance in Brazil, as the country, in addition to having a deficiency in the number of doctors per inhabitant, the number of ICU beds and ventilators available for urgent and emergency cases, presents several risk groups, such as: elderly over 60 years old, people with prognostic comorbidity, indigenous people and population’s great genetic variation (Marson; Ortega, 2020).

Government actions to mitigate the COVID-19 epidemic curve have been adopted in several countries. In Spain, according to Saez (2020), measures of social distancing led to a cases curve flattening, after the first few days the cumulative change rate in new cases decreased by an average of 3.059 percentage points daily. In Pará state, high mortality and lethality rates were reported even with social isolation, which raised the curve of new cases and directly affected hospital care due to overcrowding by infected people. On May 6, 2020, a more extreme control policy was decreed by the State of Pará government aimed at containing the pandemic advance, setting the lockdown model in the metropolitan region and neighboring municipalities.

People’s traffic control measures are fundamental in controlling the pandemic progress. According to Kucharski et al. (2020), the cases’ reduction of COVID-19 in Wuhan observed in February 2020, coincided with traveling control measures adopted in the region.

Therefore, it has been suggested that environmental factors and social control through social distancing tend to control the new cases and deaths numbers due to COVID-19. As well, the deficiency in the health system associated with the reduced number of intensive-care physicians, low numbers of mechanical ventilators, the minimum number of available beds, and the specific medication absence tend to compromise the diagnosed population care. It is questioned: What are the dynamics of new community cases and mortality records evolution due to COVID-19 in subtropical regions with temperatures between 20 and 35 °C, with social distancing by quarantine and lockdown adoption?

## 2. Materials and methods

### 2.1 Study area

The study was carried out in 114 affected municipalities among the 144 existing in the state of Pará, North region, Brazil (Fig.1). The state has a territorial extension of 247,689,515 km^2^ and an estimated population of 8,602,285 people, with 8,191,559 residents in urban areas and 2,389,492 in rural areas, with a population density of 6.07 inhabitants/km^2^ and an average Human Development Index of (HDI = 0.698) (BIGS, 2020).

**Fig. 1.**
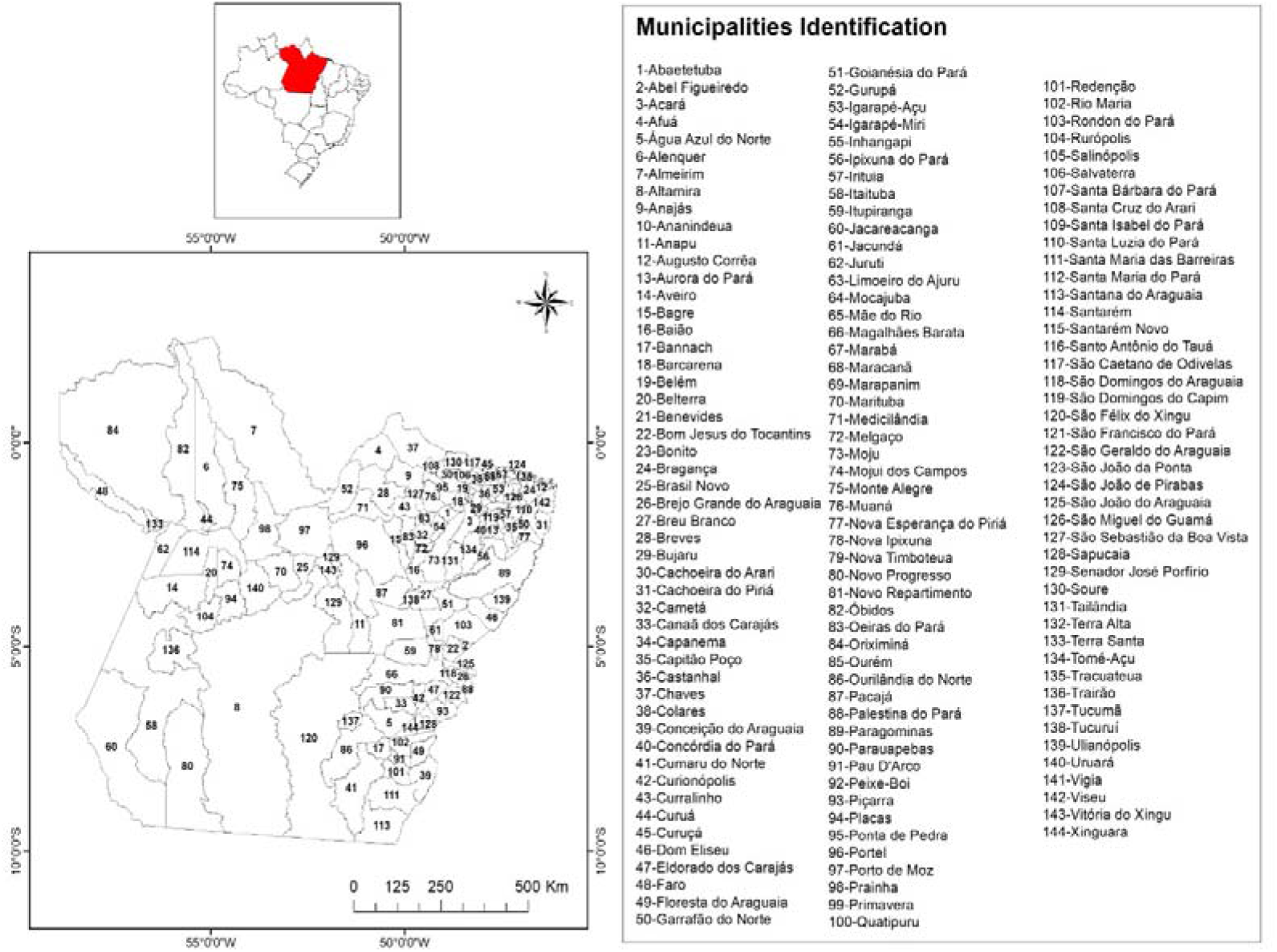
State of Pará, Northern Brazil and its 144 municipalities.

### 2.2. Data collection

The cumulative daily number of deaths by age group and sex of COVID-19 cases in 114 municipalities in the state of Pará, with a record of infection, were obtained through the daily monitoring of technical bulletins provided by the State of Pará Public Health Secretary (SHSP, 2020). General data for Brazil and its northern region were obtained from the Secretary of Health Surveillance of the Ministry of Health (BRAZIL, 2020).

### 2.3. Statistical analysis

Since understanding the dynamics of infection spread can favor more effective public control policies and allow control of transmission in new regions, an exploratory data analysis (EDA) was carried out, with numerical variables described using means, standard deviations, coefficient of variation, distributions and Pareto. Weekly maps were built for the new accumulated cases, in the period of 49 days, to assess the spread of the COVID-19 pandemic in the municipalities of the state of Pará. It was used a classification method using the Jenks algorithm based on the Absolute Deviations over the Median of the Classes.

## 3. Results and Discussion

### 3.1 Descriptive analysis

In Brazil, on May 5, 2020, 114,715 cases of COVID-19 were registered and 7,921 deaths, with a lethality rate of 6.8%, the Southeast and Northeast regions are the most affected counting around 75% of the registered cases (Fig 2B). The most affected regions in the country are the Southeast (64,756; 44.6%), followed by the Northeast (45,724; 31.5%) and North (23,207; 16.0%) (BRASIL, 2020). In the North, the state of Amazonas had the largest number of confirmed infected, 10,727 cases and disease lethality around 874 registered deaths The country on May 6, 2020, registered 114,715 cases and 7,921 deaths.

**Fig. 2.**
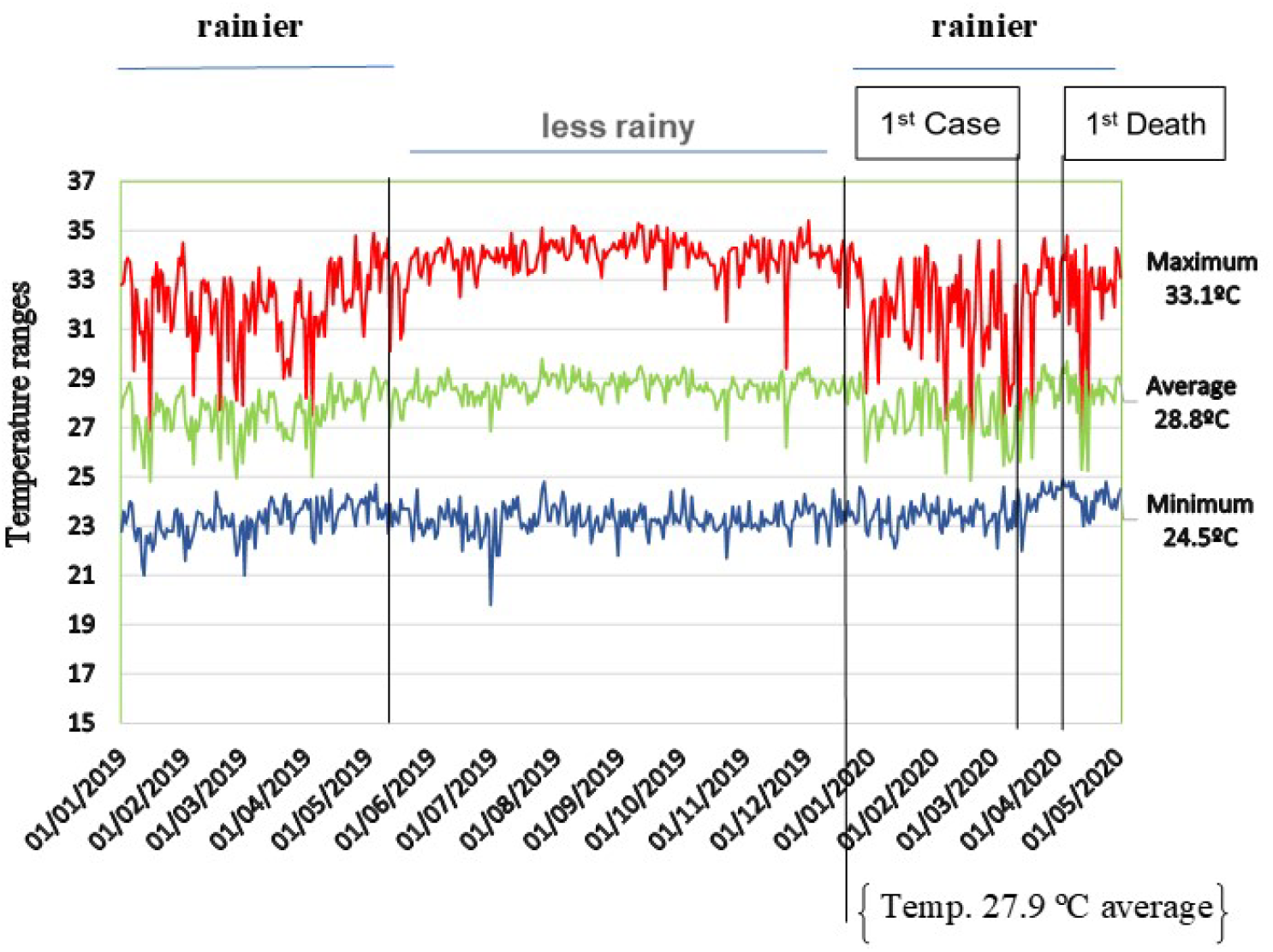
Temperature behavior (°C), rainfall behavior, 1^st^ confirmed case and 1^st^ death registration by COVID-19 in the state of Pará.

In Northern Brazil, the federative units most affected on May 5, 2020, in terms of the spatial distribution of registered cases of COVID-19 with an incidence and mortality rate per 1,000,000 inhabitants, were Amazonas (2327.5 and 251.7) and Pará (627.4 and 49.5). The capital of Pará has the highest incidence (1,816.4/1,000,000 inhabitants) Fig. 3A) and mortality (240/1,000,000 inhabitants) with a mortality rate of 9.9% (Fig. 3B).

**Fig. 3.**
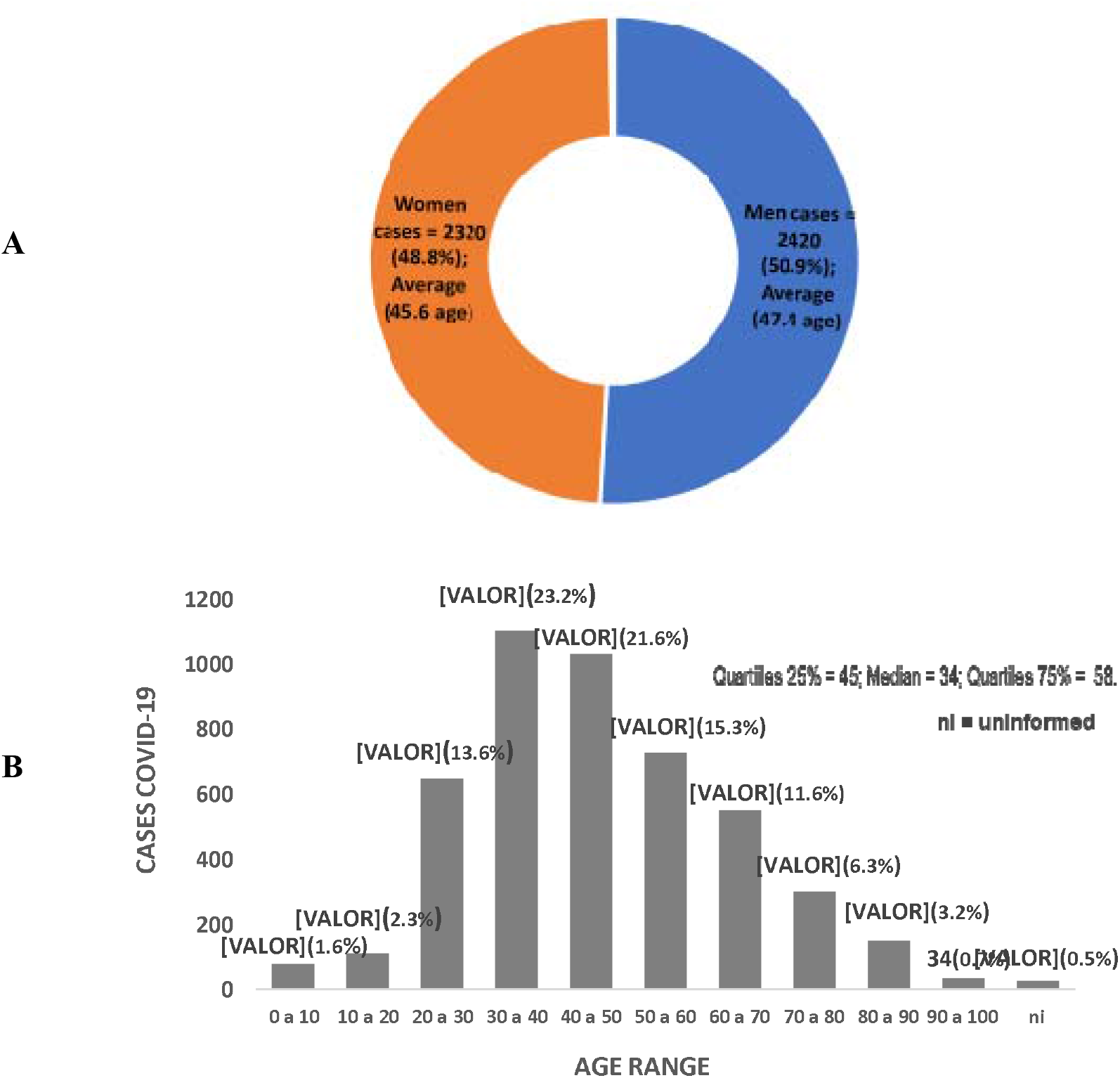
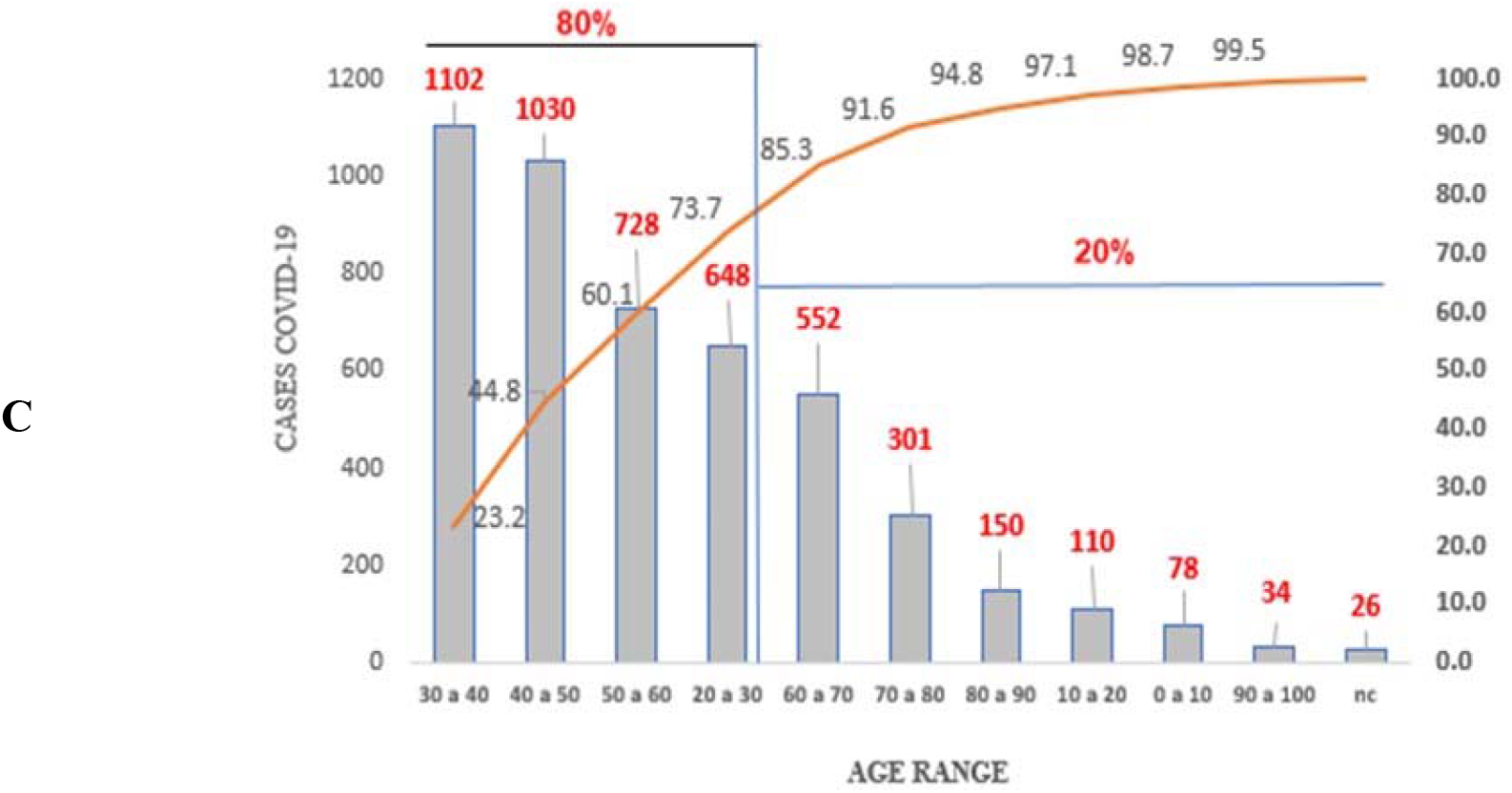
Exploratory statistical analysis of the confirmed cases accumulated of COVID-19 by sex (**A**), age group (**B**) and Pareto distribution of ages (**C**) of those infected, Pará-Brazil.

Predictive studies in Brazil report smaller records of new cases of COVID-19 at temperatures around 25.8°C, reducing the behavior of curve growth (Prata et al., 2020). Since SARS-CoV-2 may be vulnerable to fluctuations in environmental conditions similar to other coronaviruses (Le et al., 2020). It is important to note that climatological factors variability can interfere with the curve behavior even at higher temperatures.

The northern region of Brazil has an equatorial climate ranging from humid to semi-arid, with temperatures ranging from 20 to 35 °C. In the state of Pará, the temperature presents spatial and seasonal homogeneity, with an average variation of 25 °C to 35 °C. In the region, there are two distinct periods of temperature ranges, classified as rainier between December to May and less rainy between June to November, according to rainfall variation that occurs in the Amazon. The metropolitan region of Belém city in the period between January 1, 2020, to May 5, 2020, presented temperature variation between 24 °C to 31 °C and relative humidity between 70 to 80%. In this period, the number of new cases and mortality due to COVID-19 has risen since the first case notification on March 18, 2020, with the first death recorded on April 1, 2020 (Fig.2).

The North region was in 3^rd^ place in cases number with approximately 16% of the confirmed cases on May 5, 2020. In the state of Pará, the outbreak of COVID-19 registered a record of 4,756 cases with an incidence rate of 788 cases in 49 days after the first infection confirmation. The first confirmed case of the disease was notified on March 18, 2020, in Belém city, capital of the state. These records occurred when the average temperature of the region, for the period considered to be the rainiest in the region, was 27.9 °C and the air relative humidity ranging from 70 to 80%. The first notification of death occurred on April 1, 2020, and since then the curve of new cases and deaths incidence have been frequent and registered with high rates in the region.

*A priori* it is suggested that there is no inverse relationship between ambient temperature and new cases of COVID-19 in Pará in the period considered as the rainiest in the Amazon region. Therefore, there is no evidence to suppose that in regions with warm climates, low relative humidity and high rainfall variation, the cases number is lower compared to regions with moderate and/or cold climates. Similar results were obtained by Jahangiri, Jarangiri, Njafgholipour (2020) in the transmission rate evaluation of the new coronavirus in different provinces of Iran.

Fig. 3 summarizes the overall cases number compared to diagnosed population sex, the cumulative daily descriptive statistics of newly notified cases of COVID-19, and the age group distribution behavior of diagnosed population with the infection since the first disease case notified in the State of Pará.

The analysis was carried out after seven weeks of the first COVID-19 case recorded in Pará, being 4,756 cases reported between March 18 to May 5, 2020. Of the total, 2,420 (50.9%) diagnosed cases were men and had an average age of 45.6 years. The women were 2,320 (48.8%) infected cases with an average age of 47.4 years. It is reported that 15 registered cases did not present notification regarding gender and were considered as unidentified (Fig. 3A). Similar results of higher COVID-19 cases prevalence in men were obtained by Chen et al. (2020) and Nikpouraghdam et al., (2020).

In Pará, 75% were under 58 years old. Only 25% of infected cases were elderly and 1.6% of those diagnosed infected were children from 0 to 10 years old. Only 26 (0.5%) registered cases had no age identified and were considered in terms of registration as “unidentified” (Fig. 3B).

In terms of the sample of the infected population age range, negative asymmetric behavior was identified for distribution, with ages concentration around the median of 34 years old, with 80% of cases diagnosed aged between 20 and 60 years old (Fig. 3C). In Iran, the cases concentration was between 30 and 70 years old for 79.1% of COVID-19 records and approximately 39% for the elderly (Nikpouraghdam et al., 2020).

Concerned with the new cases and mortality rates, the government of Pará established a decree for lockdown in the capital and additional nine municipalities to increase the social isolation index, reducing the disease spread and reducing the new cases registration number of COVID-19.

The restrictive measure was initially focused on forcing social isolation in the municipalities of Belém, Ananindeua, Marituba, Benevides, Santa Bárbara do Pará, Santa Izabel do Pará, Castanhal, which form the Belém Metropolitan Region (BMR) and inland towns, such as Santo Antonio do Tauá, Vigia de Nazaré and Breves. The lockdown arises when the state of Pará registered a COVID-19 rate in the order of 51/100,000 inhabitants, higher than the national rate. The municipalities affected by the restriction presented rates of 75/100,000 inhabitants, higher than that registered by the state.

Despite the restrictions having focused on only 10 municipal regions, it is worth mentioning that when the lockdown was set, more than 70% of the municipalities forming the state of Pará already had notifications of COVID-19 cases.

The regional disease expansion and the high registration rates in the BMR have compromised the structure of the state’s public and private healthcare systems due to the high demand for basic care services and the high need for more complex services involving hospitalizations and intubation of patients with aggravated cases.

In the Pará state, severe cases of COVID-19 and mortality records are related to several comorbidities, the most commons are associated with heart disease, diabetes, kidney disease, pneumonia, immunodeficiency, asthma, obesity, neurological disease, hematological disease and illness hepatic; the most frequent ones being associated with heart disease and diabetes (Fig. 4).

**Fig. 4.**
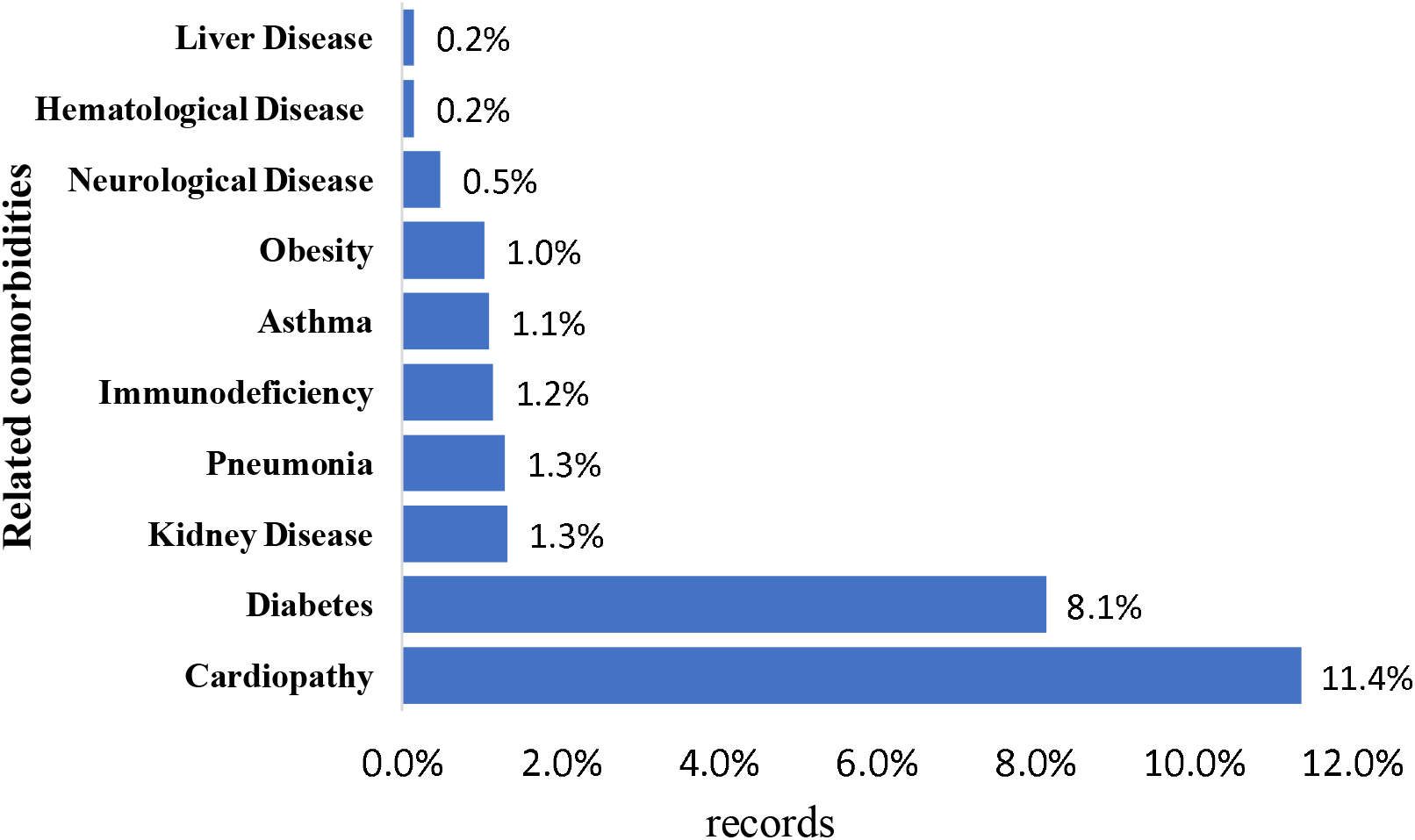
Common comorbidities in diagnosed cases of COVID-19 in the state of Pará, from March 18 to May 5, 2020.

According to Chen et al. (2020), patients infected with COVID-19 tend to have associated comorbidities with one or more chronic diseases. In Iran, patients diagnosed with COVID-19 had a strong association with chronic diseases such as diabetes, chronic respiratory diseases, hypertension, cardiovascular diseases, kidney diseases and cancer, these comorbidities were also observed in cases of deaths (Nikpouraghdam et al. 2020). Similar results were verified in infected people in Wuhan, China (Wu et al., 2020), corroborating with the results occurred in this study region.

These factors associated with the precarious service capacity of hospitals and emergency care units (ECU), the insuffcient health professionals staff, lack of rapid tests to identify infected patients, contributed to COVID-19 advance in the region. Associated with these problems, there is still an insufficient hospital beds number available according to the State Department of Public Health of Pará. There are only 249 ICU beds available for adults care in the pandemic for a population of 7,581,051 inhabitants, which represents a ratio of 1/30,445.98 bed/inhabitants, such fact leads the system to collapse in a short period during a pandemic, contributing to the disease and the number of deaths progress. Collapsing is already experienced in pediatric clinic beds (Table 1).

**Table 1:** Availability and occupation of exclusive hospital beds for COVID-19 in the public healthcare system in the state of Pará.

| Hospital bed type | Total | Available | % Occupation |
| --- | --- | --- | --- |
| Pediatric | 1109 | 497 | 55.2 |
| Pediatric clinical | 8 | 8 | 0.0 |
| adult ICU | 249 | 37 | 85.2 |
| Pediatric ICU | 7 | 4 | 42.9 |
| Intermediate unit | 26 | 25 | 3.9 |
| Neonatal ICU | 4 | 1 | 75.0 |

**Source:** Secretary of Public Health of Pará.
https://portalarquivos.saude.gov.br/images/pdf/2020/May/09/2020-05-06-BEE15-Boletim-do-COE.pdf

Due to the risk of COVID-19 infected people promoting the disease advance through community transmission to more remote regions of the state, it is essential to understand the dynamics of disease new cases to enable decision-making regarding control, prevention, treatment of infected, identify groups of risks and enable decision-making on resources allocation and allow better planning on the health system in difficult times.

Results show that a week after the first notified case, only 0.82% of the municipalities had infected people, in the second week of contagion they already had 5.56%, in the third 15.3%, in the fourth 24.3%, in the fifth 46.5%, in the sixth 69.4% and the seventh week after the first notified case, there were already 81.8% of the municipalities in the state of Pará with notified COVID-19 cases (Fig.7). The highest concentration of registered cases was in the Metropolitan Region of Belém, constituted by the capital Belém, Ananindeua, Marituba, Benevides, Santa Bárbara do Pará, Santa Izabel do Pará, Castanhal.

The measure of containment by social isolation in the region did not have an expected effect on the curve of new cases for the period under study, mainly due to the low rate of population adherence. However, the lockdown instituted on May 5, 2020, gave positive percentage results in the short-term on new cases registration in the following week of decreed intervention, with a 10.07% reduction compared to the accumulated record in the week previous epidemiologic (Fig. 5). In the long-term, the reduction of the cases may be more significant, given the greater respect and adherence of the population to isolation policies.

**Fig. 5.**
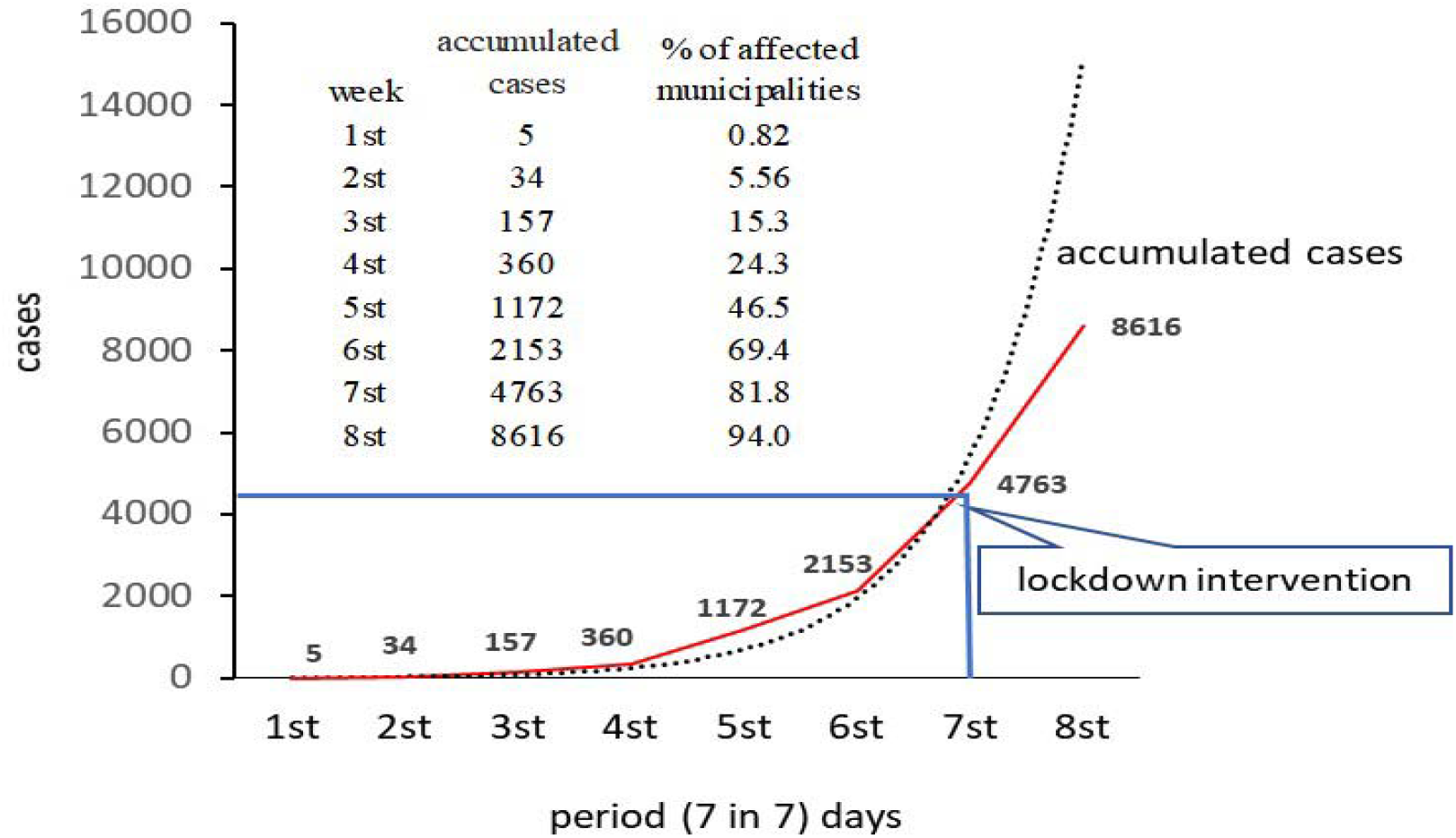
Propagation of notified COVID-19 cases in the municipalities of the state of Pará, from March 18 to May 5, 2020, and the 1^st^ post-lockdown period.

Efforts to prevent infected people from reaching more remote regions are important to stop disease transmission and prevent new cases (Coccia, 2020). To understand the dynamics of COVID-19 cases advance to remote regions from the capital, 7 maps were adjusted and show the weekly evolution of the accumulated cases registered in the municipalities of the state (Figures 6 to 12).

**Fig. 6.**
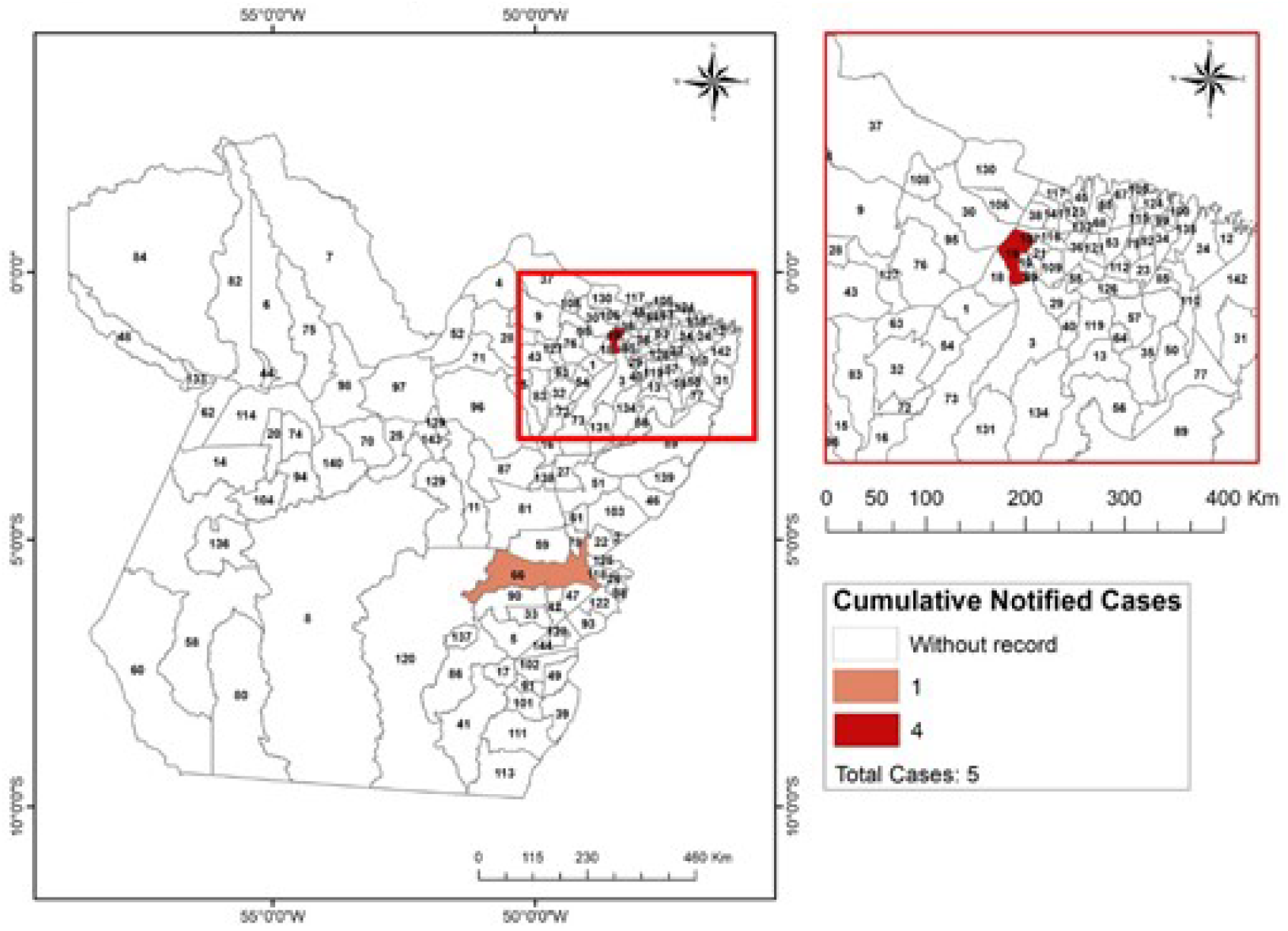
Propagation of new cases of COVID-19 accumulated in the municipalities of the state of Pará in the **1**^**st**^ **week** after registration of the first case in the region.

**Fig. 7.**
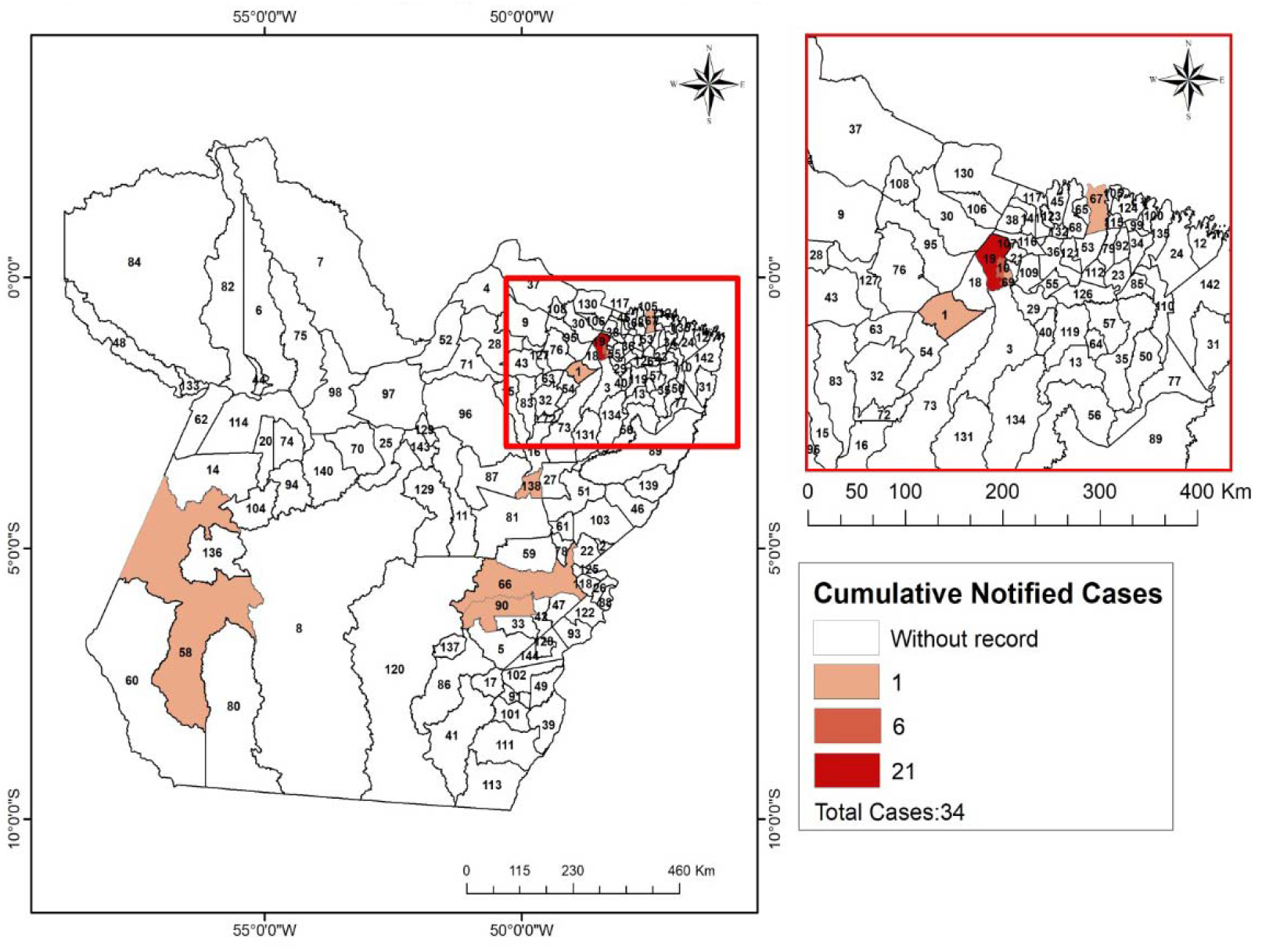
Propagation of new cases of COVID-19 accumulated in the municipalities of the state of Pará in the **2**^**nd**^ **week** after registration of the first case in the region.

**Fig. 8.**
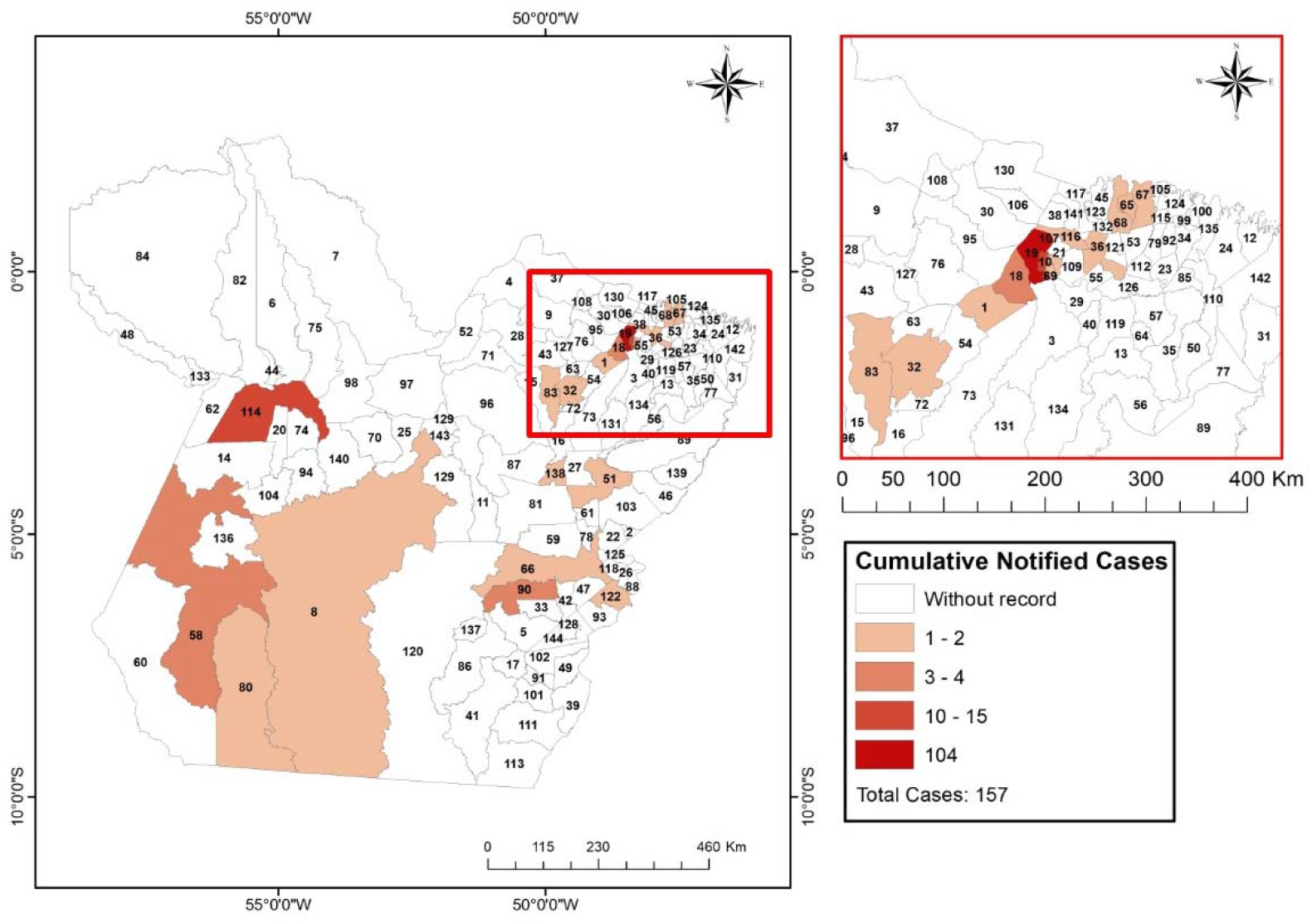
Propagation of new cases of COVID-19 accumulated in the municipalities of the state of Pará in the **3**^**rd**^ **week** after registration of the first case in the region.

**Fig. 9.**
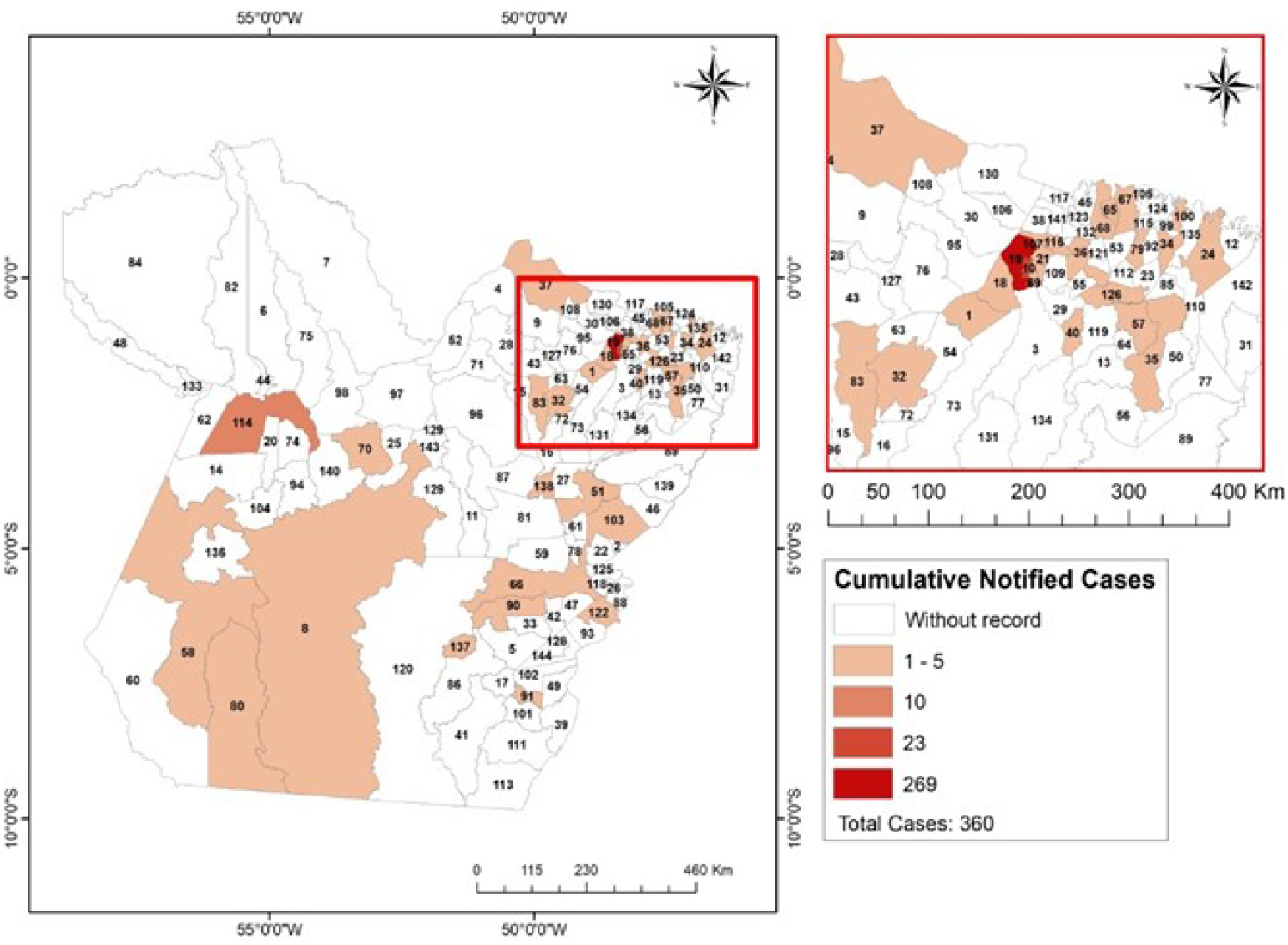
Propagation of new cases of COVID-19 accumulated in the municipalities of the state of Pará in the **4**^**th**^ **week** after registration of the first case in the region.

**Fig. 10.**
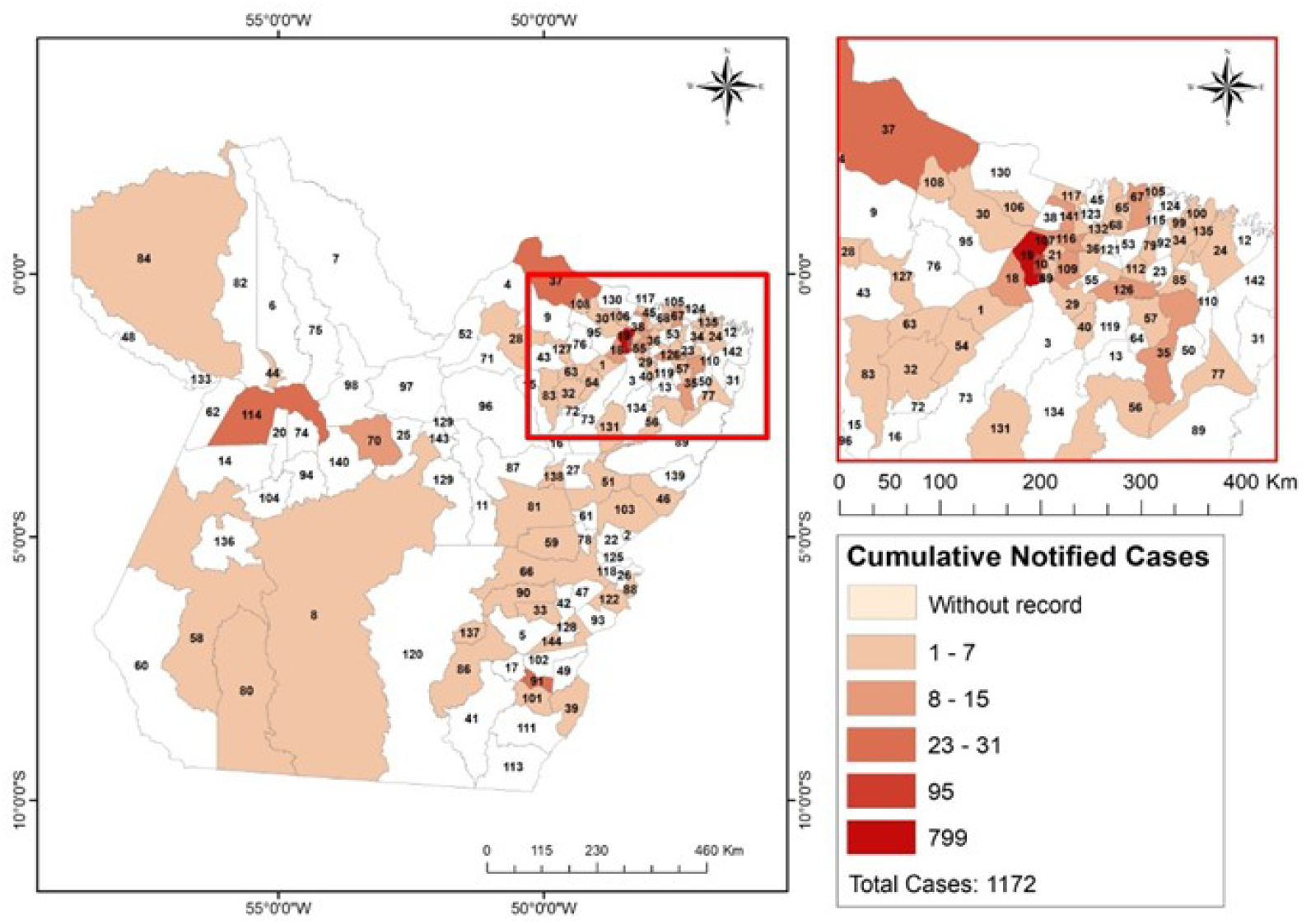
Propagation of new cases of COVID-19 accumulated in the municipalities of the state of Pará in the **5**^**th**^ **week** after registration of the first case in the region.

**Fig. 11.**
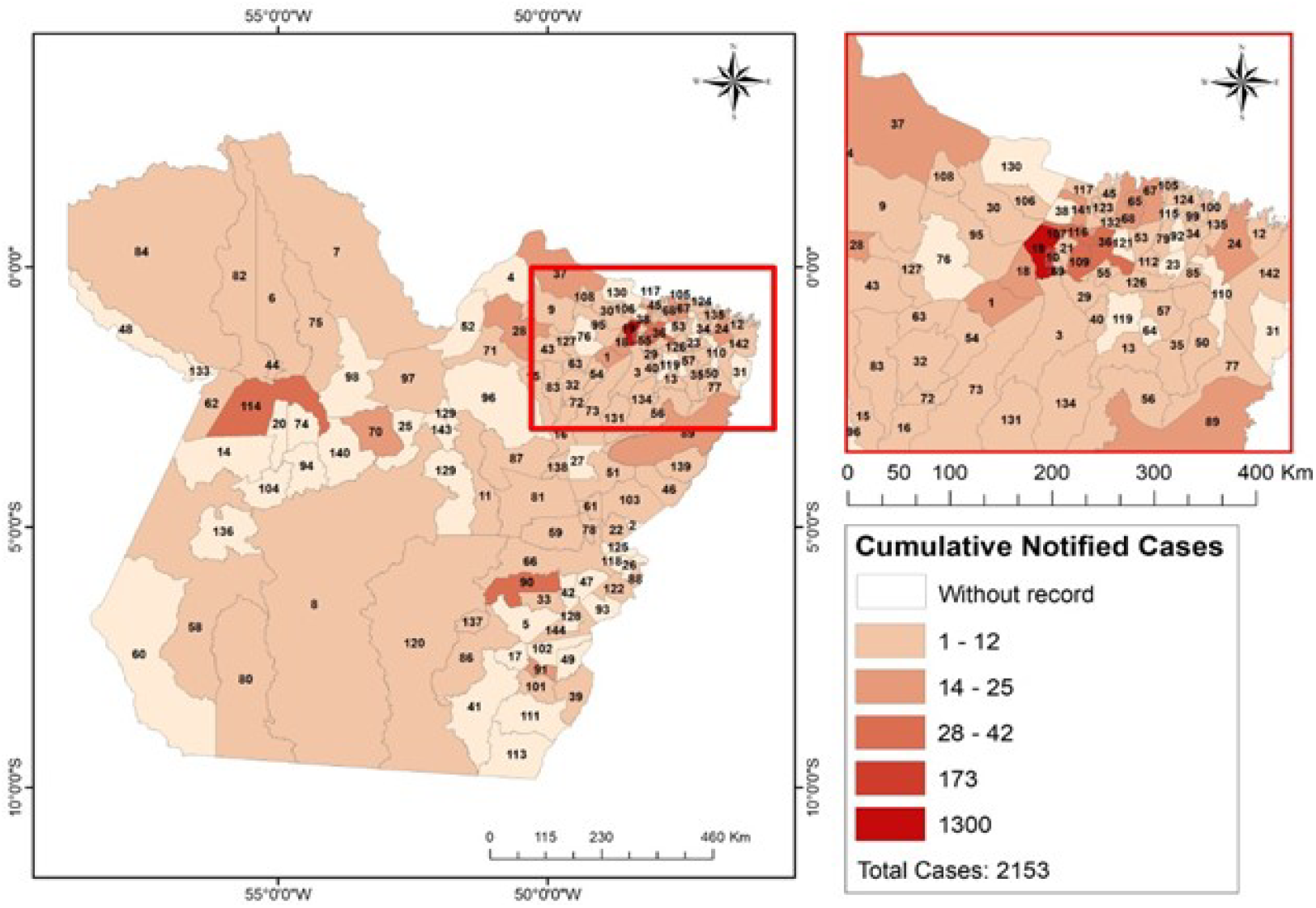
Propagation of new cases of COVID-19 accumulated in the municipalities of the state of Pará in the **6**^**th**^ **week** after registration of the first case in the region.

**Fig. 12.**
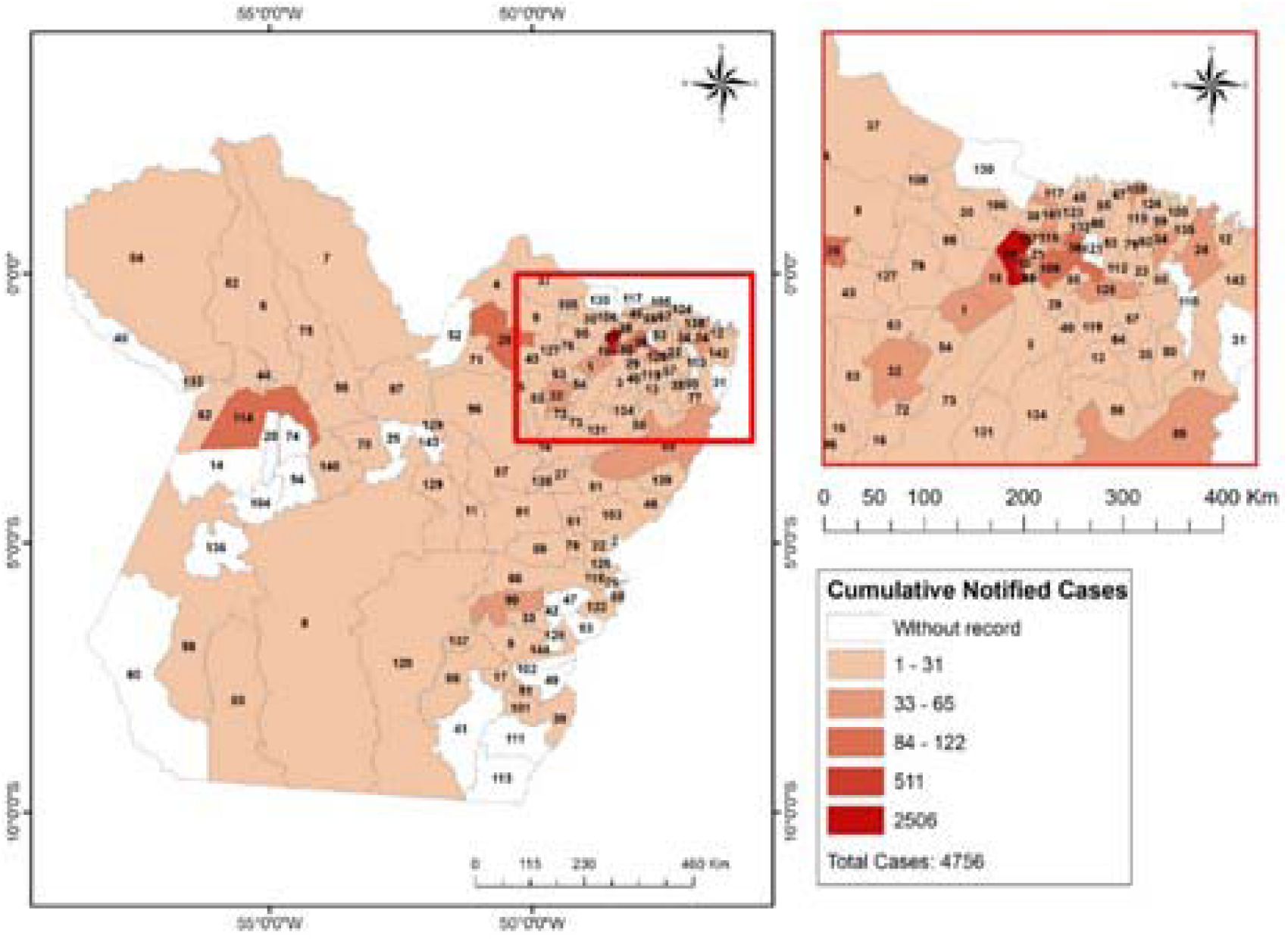
Propagation of new cases of COVID-19 accumulated in the municipalities of the state of Pará in the **7**^**th**^ **week** after registration of the first case in the region.

The municipalities that registered cases of infection in the first week were Belém (code 19) with 2 cases and Marabá (code 67) with 1 notified case (Fig. 6). The municipalities that did not present COVID-19 case records in the seven weeks after the first case were: Abel Figueiredo (code 2), Aveiro (code 14), Belterra (code 20), Brasil Novo (code 25), Brejo Grande do Araguaia (code 26), Cachoeira do Arari (code 30), Cumaru do Norte (code 41), Curionópolis (code 42), Eldourado dos Carajas (code 47), Faro (code 48), Araguaia Forest (code 49), Gurupá (code 52), Jacareacanga (code 60), Mojuí dos Campos (code 74), Piçarra (code 93), Plates (code 94), Rio Maria (code 102), Rurópolis (code 104), Santa Luzia do Pará (code 112), Santa Maria das Barreiras (code 111), Santana do Araguaia (code 113) Francisco do Pará (code 121), Sapucaia (code 128), Soure (code 130), Trairão (code 136) and Vitória do Xingu (code 120) (Fig.12).

## 4. Conclusions

Several factors can influence the spread of the virus and consequently influence the rate of incidence and mortality. In this research, the incidence rate and mortalities in Brazil and the North region were analyzed, as well as the monitoring of dynamics of the COVID-19 progress for the municipalities of the state of Pará. The results indicated that 49 days were necessary to 81% out of 144 municipalities over an area of approximately 1,248,000 km2 were affected by COVID-19 cases. Social isolation was not efficient to contain new cases advance, due to low population adherence. The social isolation and quarantine associated with the adoption of a strict measure of population circulation, the “Lockdown”, and the mandatory use of masks in public environments were effective in reducing new cases of COVID-19 registration in the short-term.

## Data Availability

We declare that all data are of a secondary nature. And they were obtained on public access platforms. According available links.

https://covid.saude.gov.br/

https://coronavirus.saude.gov.br/sobre-a-doenca#o-que-e-covid

http://www.saude.pa.gov.br/

## Conflicts of Interest Statement

The authors declare no conflicts of interest

## Acknowledgments

This study was prepared with support from the Research Group for Biosystems Management, Modeling and Experimentation - GEMAbio.

## Notes

### Competing Interest Statement

The authors have declared no competing interest.

### Funding Statement

The research received no investment for start, progress and conclusion.

### Author Declarations

Data of a secondary nature. There was no need for IRB and / or ethics committee.

